# Obesity-Related Aldosteronism is Associated with Adverse Cardiac Structure, Function, and Adiposity

**DOI:** 10.1101/2025.11.12.25340075

**Authors:** Cheng-Hsuan Tsai, Justin M. Chan, Julia Milks, Arnaldo Ferrebus, Isabelle Hanna, Sanan Mahrokhian, Andrew J. Newman, Parisien-La Salle Stéfanie, Isabela Reis Marques, Kristen Foote, Gail K. Adler, Raymond Y. Kwong, Michael Jerosch-Herold, Bertram Pitt, Anand Vaidya, Jenifer M. Brown

## Abstract

**Background:** Obesity-related aldosteronism may increase risk for adverse cardiometabolic outcomes. We investigated the association between dysregulated aldosterone production and cardiac structure, function, and adiposity under controlled physiological conditions in obese hypertensive adults.

**Methods:** Community-dwelling participants with overweight or obesity and stage 1-2 hypertension were prospectively enrolled to undergo comprehensive phenotyping of aldosterone production, along with cardiac and abdominal MRI. Aldosterone production was assessed via four controlled physiological maneuvers designed to assess both renin-independent aldosterone production: seated saline suppression testing (SST) and oral sodium loading (OSLT), and ACTH-modifiable aldosterone production: overnight dexamethasone suppression (DST), and adrenocorticotropic hormone (ACTH) stimulation. Cardiac structure and function, cardiac fat volume, hepatic fat content, and visceral-to-subcutaneous fat ratio were assessed by MRI.

**Results:** 72 participants were enrolled, with a mean age of 55.2±9.5 years, a mean BMI of 37.8±5.3 kg/m², and of whom 68.1% were women. After SST, a continuum of non-suppressible and dysregulated aldosterone production was observed, with 29.2% of participants meeting criteria for overt primary aldosteronism. Greater post-SST aldosterone levels were independently associated with greater left ventricular mass index (p<0.001), left ventricular global longitudinal strain (p=0.038), cardiac fat volume (p=0.023), and visceral-to-subcutaneous fat ratio (p=0.003). These associations between dysregulated aldosterone production and cardiac remodeling and adipose-tissue parameters were consistently replicated under conditions of oral sodium loading and dexamethasone suppression and ACTH-stimulation.

**Conclusions:** In obese adults with hypertension, dysregulated aldosterone production and overt primary aldosteronism are prevalent and independently associated with adverse cardiac remodeling and increased cardiometabolic adipose tissue volume. These findings highlight a potential pathophysiologic link between aldosterone excess and obesity-related cardiometabolic disease that should be investigated in interventional studies.

## Introduction

Obesity and hypertension are globally prevalent and interrelated cardiometabolic disorders, significantly contributing to cardiovascular morbidity and mortality^1–4^. Additionally, obesity has been increasingly recognized to promote aldosterone excess, which exerts detrimental effects on cardiometabolic systems^5–8^. Significant hemodynamic alterations are known to occur in obesity, including expanded plasma volume, elevated cardiac output, and sympathetic system activation, all of which contribute to hypertension and may modulate aldosterone production^9–12^. Beyond these hemodynamic effects, emerging evidence also suggests that adipocyte-derived factors such as leptin, oxidized fatty acids, and pro-inflammatory cytokines can stimulate aldosterone production independently of the renin–angiotensin–aldosterone system, contributing to inappropriately elevated or autonomous aldosterone production in obese individuals^6, 7, 13–17^. Therefore, this adipose–aldosterone interaction may further exacerbate adipocyte dysfunction, promote sodium retention and myocardial fibrosis, and drive adverse cardiac remodeling, suggesting that aldosteronism may represent a mechanistic link between obesity and cardiometabolic injury^18^.

Despite these mechanistic insights, the prevalence, determinants, and implications of dysregulated aldosterone production in mediating changes in cardiac structure, function, and cardiometabolic fat accumulation in obese hypertensive individuals remains poorly understood. In this study, we conducted comprehensive physiological phenotyping of aldosterone dysregulation alongside high-resolution cardiac and abdominal magnetic resonance imaging (MRI) to elucidate the pathophysiological links between obesity, aldosteronism, and subclinical cardiovascular remodeling in obese adults without overt cardiovascular disease.

## Methods

### Data Availability

Supporting data, statistical code, and study protocols can be made available to other investigators on reasonable request to the corresponding author.

### General Overview of Study Design

Obese or overweight individuals with metabolic risk factors and mild hypertension (0-1 antihypertensives) were prospectively enrolled in a mechanistic clinical trial evaluating the impact of mineralocorticoid receptor blockade on cardiac MRI parameters at Brigham and Women’s Hospital, Boston, MA (clinicaltrials.gov NCT04519164). The analysis reported herein aimed to investigate the cross-sectional relationship of aldosterone production to baseline cardiac structure and function and cardiometabolic fat in the study population. Among the 80 obese hypertensive participants enrolled in the parent clinical trial, 72 completed the saline suppression test (SST) and were included in the present analysis. All included participants underwent four controlled physiological phenotyping maneuvers including the SST, and cardiac MRI (**Figure 1**).

**Figure 1.**
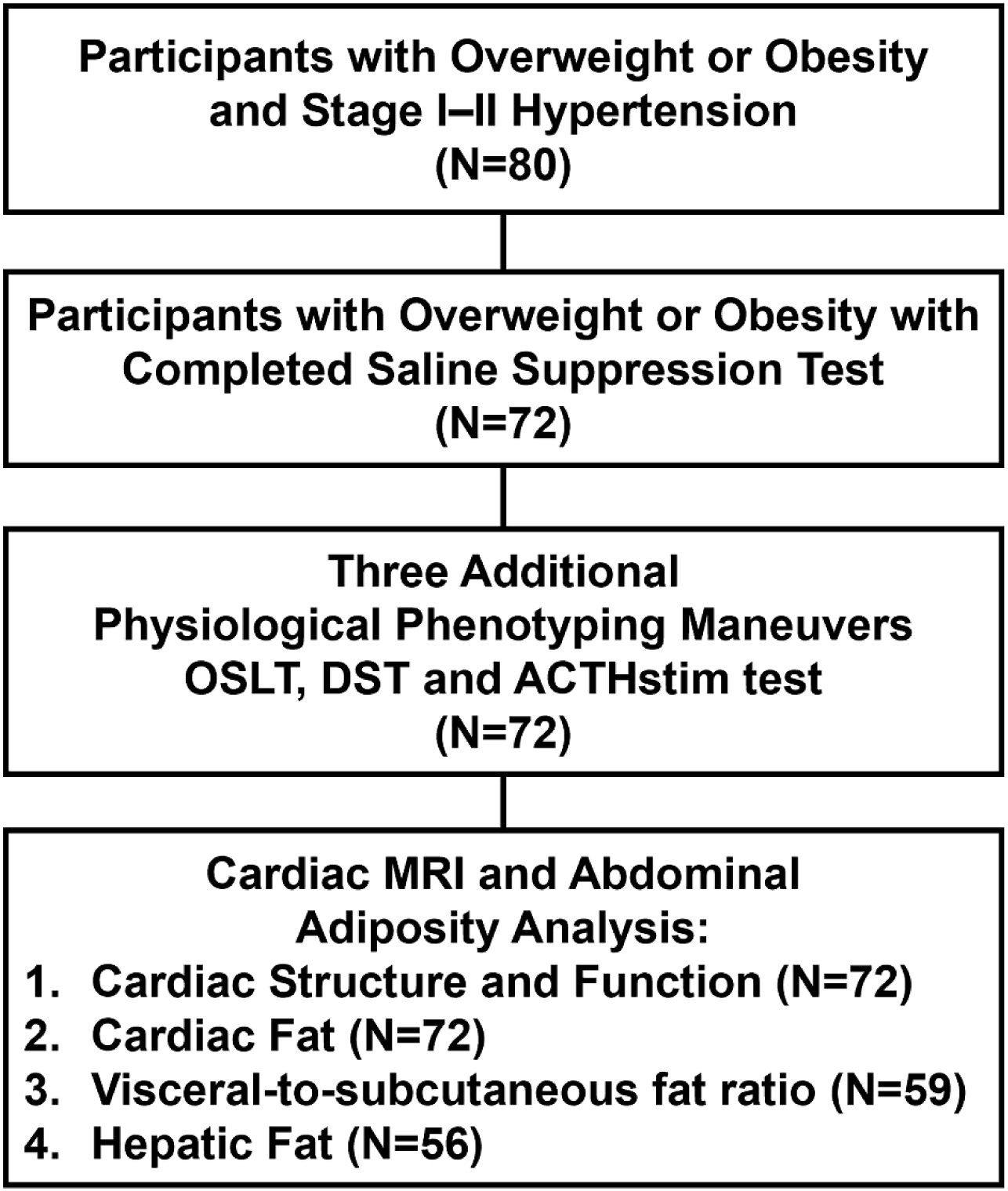
Flowchart for Assessment of Dysregulated Aldosterone Production, Cardiac Parameters, and Abdominal Adiposity in Hypertensive Participants with Overweight or Obesity. **Abbreviation:** ACTHstim test= adrenocorticotropic hormone stimulation test; DST = dexamethasone suppression test; OSLT = oral salt loading test

The SST was used as a physiologic maneuver to investigate non-suppressible, renin-independent aldosterone production. Additional detailed physiological maneuvers were performed to evaluate the consistency of the findings across phenotypes of aldosteronism, including an oral salt loading test (OSLT) to assess renin-independent aldosterone production, as well as an overnight 1 mg dexamethasone suppression test (DST) and a 250 mcg adrenocorticotropic hormone (ACTH) stimulation test to assess ACTH-modifiable aldosterone production. Together, these independent tests allowed for comprehensive phenotyping of aldosterone regulation by interrogating both the renin-angiotensin and ACTH axes. Cardiac MRI was used to measure cardiac fat, cardiac structure, function and myocardial perfusion; liver fat content and visceral-to-subcutaneous fat ratio were evaluated from additional abdominal MRI sequences included with the cardiac MRI acquisition (**Figure 1**).

### Study Population

Study participants were recruited from the local Boston, MA metropolitan area using a combination of social media advertisements and patient portal research invitations. The target population was overweight to obese with hypertension and elevated metabolic risk to enhance the likelihood of dysregulated aldosterone production and mineralocorticoid receptor activation, though no participants had been clinically diagnosed with primary aldosteronism. Participants were eligible for inclusion if they met one of two primary criteria: body mass index (BMI) ≥ 30 kg/m² with at least one of the following comorbidities, or BMI ≥ 25 kg/m² with at least two of the following comorbidities: (1) stage I (blood pressure: 120–139/80–89 mmHg) or stage II (blood pressure: 140–159/90–99 mmHg) hypertension, managed with zero to one anti-hypertensive agent; (2) dysglycemia (impaired fasting glucose 100–125 mg/dL or glycated hemoglobin 5.7–6.4%); or (3) dyslipidemia (fasting triglycerides >150 mg/dL and HDL cholesterol <40 mg/dL in men or <50 mg/dL in women).

Exclusion criteria included: (1) estimated glomerular filtration rate (eGFR) < 60 mL/min/1.73m²; (2) diagnosed or currently treated type 1 or type 2 diabetes; (3) history of cardiovascular disease (e.g., myocardial infarction, heart failure, atrial fibrillation, stroke); (4) electrocardiographic evidence of ischemia (ST-segment or T-wave changes, Q waves in more than one territorial lead, or left bundle branch block); (5) current pregnancy or breastfeeding; or (6) presence of MRI-incompatible medical implants or devices; (6) systolic blood pressure ≥ 160 mmHg, diastolic blood pressure ≥ 100 mmHg, or hypertension requiring ≥ 2 antihypertensive agents for control. All study participants provided written informed consent prior to participation, and the study protocol was approved and monitored by the Mass General Brigham Institutional Review Board.

### Physiological Characterization of Aldosterone Production

Participants underwent controlled physiological profiling to assess aldosterone production under the regulatory influence of both renin/angiotensin II and ACTH. Interfering medications that modulate the renin-angiotensin system including diuretics, ACE inhibitors, and angiotensin receptor blockers were withdrawn two weeks prior to phenotyping procedures. During the washout period, if blood pressure exceeded 130/80 mmHg at the time of medication withdrawal, doxazosin (2–8 mg/day) and/or amlodipine (2.5–10 mg/day) were initiated to maintain control. The target blood pressure range during this period was 105–150/50–90 mmHg.

To evaluate the degree of dysregulated aldosterone production, all participants underwent a seated SST between 8:00 and 9:00 AM. They were seated upright in an exam room for 30 minutes prior to a baseline blood draw, then remained seated for 4 hours while receiving a 2-liter intravenous infusion of normal saline. Plasma renin activity and plasma aldosterone concentrations were measured at the beginning and at the end of the infusion. Participants separately completed an OSLT, which involved a high-sodium diet (≥200 mmol/day) with 50 to 75 mmol/day of potassium for 4 to 7 days. Plasma aldosterone was measured at the completion of the diet at 8:00 to 10:00 am following 30 minutes of seated posture.

At a separate study visit, approximately 4-7 days later, participants underwent both a DST and an ACTH stimulation test to assess ACTH-modifiable aldosterone production. For the DST, participants took a 1-mg dose of dexamethasone between 11:00 PM and midnight, and fasting blood samples were collected the following morning between 8:00 and 9:00 AM. Immediately thereafter, a 250 mcg intravenous bolus of cosyntropin (synthetic ACTH) was administered, with an additional blood sample drawn one hour later. Plasma aldosterone levels were measured following both the DST and the ACTH stimulation test.^19^

### Laboratory Assays

Plasma aldosterone concentration (PAC) was measured using a commercially available ELISA (IBL-America; RRID: AB_2813725), with an inter-assay coefficient of variation (CV) of 8.6%–9.4%, intra-assay CV of 3.9%–9.7%, analytical sensitivity <5.7 pg/mL, and a dynamic range of 5.7–1000 pg/mL. Plasma renin activity was measured by ELISA (IBL-America, Minneapolis, MN, USA; RRID: AB_3532145), with inter-assay CV of 4.8%–7.1% and intra-assay CV of 6.3%–8.7%.

### 24-hr Ambulatory Blood Pressure Measurement

24-hour ambulatory blood pressure monitoring (Microlife WatchBP O3 Ambulatory; Microlife USA, Clearwater, FL) was performed following the washout, between the phenotyping visits, with automated measurements from 06:00 to 07:00.

### Cardiac MRI

Myocardial characteristics, cardiac structure, and cardiac function were analyzed using contrast-enhanced rest/stress MRI. All examinations were performed using a 3-T system (Prisma Fit, Siemens Healthineers, Erlangen) at Brigham and Women’s Hospital, as previously described^19–21^. Left ventricular (LV) structure by mass, end-diastolic volume and end-systolic volume and LV function by stroke volume and ejection fraction (EF) were measured by blinded staff of the Brigham and Women’s cardiac MRI core lab (Mass Medis, Medis Medical System, Leiden)^22^. We additionally evaluated systolic function by LV mid-wall global longitudinal strain and global circumferential strain by feature tracking on cine sequences^19^. Indexed parameters were divided by body surface area. Perfusion images were analyzed with QMASS CMR image analysis software by tracing the endocardial and epicardial contours, to generate signal intensity vs. time curves to analyze myocardial contrast enhancement. The LV wall was segmented into 6 sectors. A region of interest was placed in the blood pool to obtain an arterial input for perfusion analysis, respectively.

Extracellular volume (ECV) fraction was calculated by plotting the reciprocal of segmental myocardial T1 values against the reciprocal of blood pool T1 values from native, 10-minute, and 20-minute postcontrast modified Look-Locker inversion recovery acquisitions. Linear regression slopes from these plots, representing the segmental partition coefficient for gadolinium, were multiplied by (1 − hematocrit) to derive the segmental ECV. The global LV ECV fraction was obtained by averaging the segmental ECV values^23^. Cardiac MRI myocardial perfusion imaging was performed at rest and during pharmacologic stress. Stress was induced using intravenous regadenoson infusion (400 microgram bolus administered through an intravenous line, followed by flush) prior to image acquisition. A gadolinium-based contrast agent (Gadobutrol, Gadavist; 0.05 mmol/kg) was administered at a rate of 4-5 mL/s. First-pass perfusion imaging using a T1-weighted saturation recovery gradient-echo sequence was obtained in 2–3 short-axis slices under both rest and stress conditions, with approximately 15 minutes between the two acquisitions. Weight-based IV aminophylline was used to reverse regadenoson immediately after stress perfusion. Global myocardial blood flow was calculated as the average across all segments and slices. Signal intensity curves from the perfusion images were converted to R1 (T1 relaxation rate) vs. time curves, using model-based calibration as described previously^24^. Rest and stress myocardial blood flows were quantified from segmental R1 vs. time curves by model-independent deconvolution^25^. Myocardial perfusion reserve was defined as the ratio of stress to rest myocardial blood flow^26^.

### Cardiometabolic adiposity measurement

Cardiac fat volume was measured from the cardiac MRI using the T1-weighted sequences (**Figure 2A**). This measurement included both epicardial fat (within the pericardium) and pericardial fat (outside the pericardium). Due to the strong correlations of both pericardial and epicardial fat with total cardiac fat in our cohort (r = 0.89 and 0.86, respectively; p < 0.001), along with the lower reproducibility of epicardial fat quantification, we chose to evaluate total cardiac fat volume. Fat segmentation was performed using anatomic landmarks to isolate the heart and pericardial fat from the thorax. The anterior boundary was the chest wall, and the posterior boundary was the aorta and bronchus. Manual regions of interest (ROIs) were drawn encompassing the cardiac fat on each slice, and total cardiac fat volume was calculated as the sum of voxels containing fat^20, 27^.

**Figure 2.**
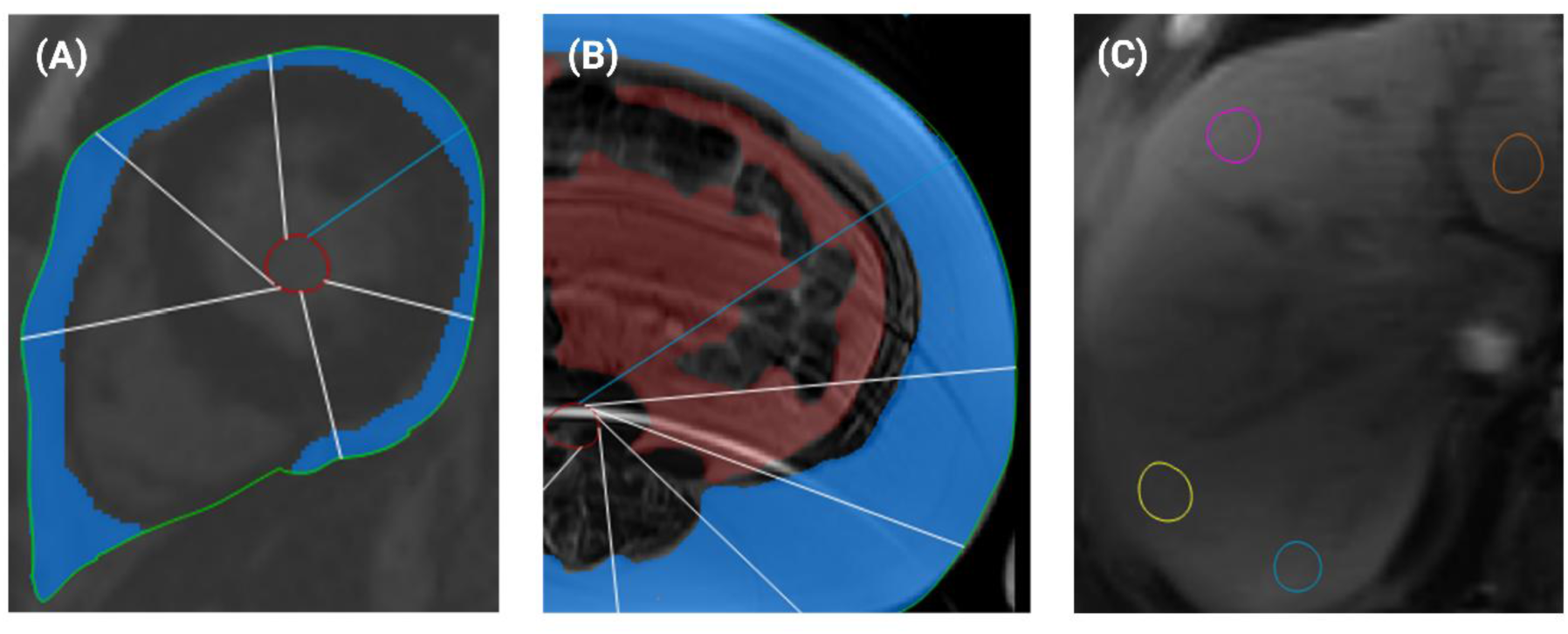
MRI-Based Quantification of Cardiometabolic Adiposity. (A) Cardiac fat volume was quantified on cardiac MRI using T1-weighted sequences and included both epicardial and pericardial fat. (B) Visceral adiposity was defined as fat enclosed within the visceral cavity; subcutaneous adiposity was defined as fat outside the visceral cavity, excluding fat within the muscular fascia, on T1-weighted images. (C) Hepatic fat content was measured with a chemical-shift–encoded Dixon technique to generate PDFF maps; regions of interest were placed in four hepatic lobes (right anterior, right posterior, left medial, left lateral).

During the cardiac MRI described above, dedicated abdominal imaging sequences were also acquired. Abdominal adiposity was assessed from an axial slice at the level of the L3–L4 intervertebral disc. Visceral adiposity was defined as fat enclosed within the visceral cavity, while subcutaneous adiposity was defined as fat located outside the visceral cavity but excluding fat within the muscular fascia^28^. T1-weighted MRI sequences were used to manually delineate fat boundaries (**Figure 2B**). The visceral-to-subcutaneous fat ratio was calculated as an index of visceral obesity^29^.

Hepatic fat content was quantified using a chemical shift–encoded MRI technique based on the Dixon method to generate proton density fat fraction (PDFF) maps, yielding a percentage of fat over fat plus water content. ROIs were manually placed in four different hepatic lobes (right anterior, right posterior, left medial, and left lateral) on PDFF maps, carefully avoiding large vessels, bile ducts, and artifacts (**Figure 2C**). The mean value of these four ROIs was used to represent the hepatic fat content^30^.

Although all enrolled participants completed cardiac MRI assessments, due to inadequate image quality, visceral-to-subcutaneous fat ratio and hepatic fat could be measured in only 59 participants and 56 participants, respectively. Quantification of cardiometabolic fat, including cardiac, visceral, subcutaneous, and hepatic fat, was performed by independently by three readers, C.H.T., J.M.B., and J.C. The method demonstrated excellent inter-reader reliability, with intraclass correlation coefficients (ICCs) greater than 0.97 for all fat depots measured. All image analyses were performed blinded to hormonal data.

### Statistical Analysis

Data are presented as mean ± standard deviation for normally distributed variables, and as median (interquartile range, IQR: 25th and 75th percentiles) for non-normally distributed variables. The Kolmogorov–Smirnov test was used to assess normality of continuous variables. Analyses were conducted using complete-case data; participants with missing values for any covariates were excluded from the corresponding models. Waterfall plots were generated to depict the raw distribution of dysregulated aldosterone levels after the SST and to visualize the continuum of dysregulated aldosterone production across study participants. To assess measurement reproducibility, ICCs were calculated using a two-way random-effects model with absolute agreement. Pearson correlation coefficients (r) were used to evaluate the associations between total cardiac fat volume and both epicardial and pericardial fat volumes. To evaluate the associations between aldosterone measurements under four physiological conditions: SST, OSLT, DST and ACTH stimulation test, and the cardiac MRI-derived structural, functional, and cardiometabolic fat parameters, univariable and multivariable linear regression analyses were performed. Multivariable models were adjusted for age, sex, BMI, 24-hour ambulatory systolic blood pressure, and eGFR. A sensitivity analysis was conducted among participants with complete visceral-to-subcutaneous fat ratio measurements. Aldosterone concentrations under each physiologic condition were log-transformed to normalize their distribution.

To account for the consistency of repeated measurements of aldosterone obtained under the four physiological conditions in each participant, linear mixed-effects models were applied. A random intercept was included for each subject to account for intra-individual correlation. The primary fixed effects, including LVMI, LV global longitudinal strain, cardiac fat volume, and the ratio of visceral-to-subcutaneous fat, were modeled for their associations with log-transformed plasma aldosterone concentrations. Multivariable models were again adjusted for age, sex, BMI, 24-hour ambulatory systolic blood pressure and eGFR. Model parameters were estimated using restricted maximum likelihood. The overall significance of each fixed effect across conditions was evaluated using a global p-value derived from the mixed-effects model.

An exploratory analysis based on baseline renin was conducted to examine the potential influence of renin-dependent versus renin-independent aldosterone production. Waterfall plots were generated for participants with low baseline plasma renin activity (≤1.0 ng/mL/h). Additionally, low baseline renin status (plasma renin activity ≤1 ng/mL per hour) was included as an additional covariate in the multivariable linear regression and linear mixed-effects models to evaluate whether the relationships with aldosteronism were driven by renin. Statistical significance was defined as two-sided p < 0.05. Analyses were performed using SPSS for Windows (version 25.0; SPSS, Inc., Chicago, IL) and Python (version 3.11) with the statsmodels package (version 0.14.0).

## Results

### Characteristics of study participants

Baseline characteristics of the study participants are summarized in Table 1. The mean age was 55.2±9.5 years, and 68.1% of the participants were women. The mean BMI was 37.8±5.3 kg/m^2^. The screening seated blood pressure was 129.1 ± 11.5 mmHg systolic and 82.0 ± 8.6 mmHg diastolic, measured while 47 of 72 participants (65.3%) were taking a single antihypertensive medication with the remainder untreated. During the washout period when diuretics, ACE inhibitors, and angiotensin receptor blockers were withdrawn, the mean 24-hour ambulatory blood pressure was 124.4 ± 8.8 mmHg systolic and 73.7 ± 6.9 mmHg diastolic. The median plasma renin activity was 1.3 (IQR: 0.5, 2.5) ng/mL/h, and the median PAC was 18.8 (IQR: 14.8, 27.0) ng/dL at baseline prior to SST.

**Table 1.** Clinical characteristics.

|  | Participants (N=72) |
| --- | --- |
| **Clinical characteristics** |  |
| Age, year | 55.2±9.5 |
| Sex, women, % | 49 (68.1) |
| Body mass index, kg/m<sup>2</sup> | 37.8±5.3 |
| Screening SBP, mmHg | 129.1±11.5 |
| Screening DBP, mmHg | 82.0±8.6 |
| 24hr SBP, mmHg | 124.4±8.8 |
| 24hr DBP, mmHg | 73.7±6.9 |
| eGFR, ml/min/1.73m<sup>2</sup> | 87.5±14.2 |
| Potassium, mmol/L | 4.3±0.4 |
| Hemoglobin A1C, % | 5.3±0.4 |
| Low-density lipoprotein, mg/dL | 115.4±27.9 |
| Triglycerides, mg/dL | 139.5±66.4 |
| Plasma renin activity, ng/mL per hour | 1.3 (0.5, 2.5) |
| PAC, ng/dL | 18.8 (14.8, 27.0) |
| ARR, ng/dL per ng/ mL per hour | 14.7 (12.7, 18.9) |
| **Physiological phenotyping** |  |
| Saline suppression test PAC, ng/dL | 11.5 (7.7, 14.1) |
| Oral salt loading test PAC, ng/dL | 15.7 (10.0, 22.8) |
| Dexamethasone suppression test PAC, ng/dL | 13.3 (8.8, 19.7) |
| ACTH-stimulation test PAC, ng/dL | 38.0 (24.4, 52.1) |
| **Cardiac MRI** |  |
| LV ejection fraction, % | 59.4±5.6 |
| LV end-systolic volume index, mL/m<sup>2</sup> | 29.0±6.4 |
| LV end-diastolic volume index, mL/m<sup>2</sup> | 71.4±13.3 |
| LV mass, g | 98.4±24.8 |
| LV mass index, g/m<sup>2</sup> | 49.1±10.6 |
| LV global longitudinal strain, % | -18.6±3.4 |
| LV global circumferential strain, % | -21.9±3.7 |
| Left atrial volume index, mL/m<sup>2</sup> | 42.0±12.0 |
| Left atrial ejection fraction, % | 54.8±10.5 |
| Left atrial strain, % | 34.3±10.3 |
| RV end-systolic volume index, mL/m<sup>2</sup> | 33.5±8.5 |
| RV end-diastolic volume index, mL/m<sup>2</sup> | 71.8±14.2 |
| LV extracellular volume fraction | 27.6±2.3 |
| Resting Myocardial Blood Flow, ml/min/g | 1.0±0.2 |
| Stress Myocardial Blood Flow, ml/min/g | 2.5±0.6 |
| Myocardial perfusion index | 2.4±0.6 |
| **Cardiac and abdominal fat assessments\*** |  |
| Cardiac fat volume, ml | 217.5±75.2 |
| Subcutaneous fat area*, cm<sup>2</sup> | 311.0±134.0 |
| Visceral fat area*, cm<sup>2</sup> | 240.6±93.8 |
| Visceral-to-subcutaneous fat ratio* | 0.9±0.5 |
| Hepatic fat*, % | 17.2±11.4 |

\*Subcutaneous fat area, visceral fat area, and the visceral-to-subcutaneous fat ratio were measured in 59 participants, while hepatic fat content was measured in 56 participants.
**Abbreviations:** ACTH = adrenocorticotrophic hormone; ARR = aldosterone-to-renin ratio; DBP = diastolic blood pressure; eGFR = estimated glomerular filtration rate; LV = left ventricular; MRI = magnetic resonance imaging; PAC = plasma aldosterone concentration; SBP = systolic blood pressure; RV = right ventricular.

### Physiological phenotyping

There was a broad spectrum of dysregulated aldosterone production following SST, ranging from complete physiological suppression to overtly non-suppressible production (**Figure 3A**), with a median post-SST PAC of 11.5 ng/dL (interquartile range: 7.7–14.1 ng/dL). When applying the 2025 Endocrine Society Clinical Practice Guideline definition of primary aldosteronism using an SST (baseline PRA ≤ 1 ng/mL/h and post-SST PAC ≥ 7.8 ng/dL by immunoassay) ^31^, 21 of 72 participants (29.2%) met criteria for overt primary aldosteronism. An additional 13 of 72 participants (18.1%) exhibited subclinical primary aldosteronism (baseline PRA ≤ 1 ng/mL/h and post-SST PAC < 7.8 ng/dL by immunoassay), with the remainder demonstrating renin-dependent, non-suppressible aldosterone production (**Figure 3**). Mean aldosterone levels measured under the four phenotyping conditions are summarized in **Table 1**.

**Figure 3.**
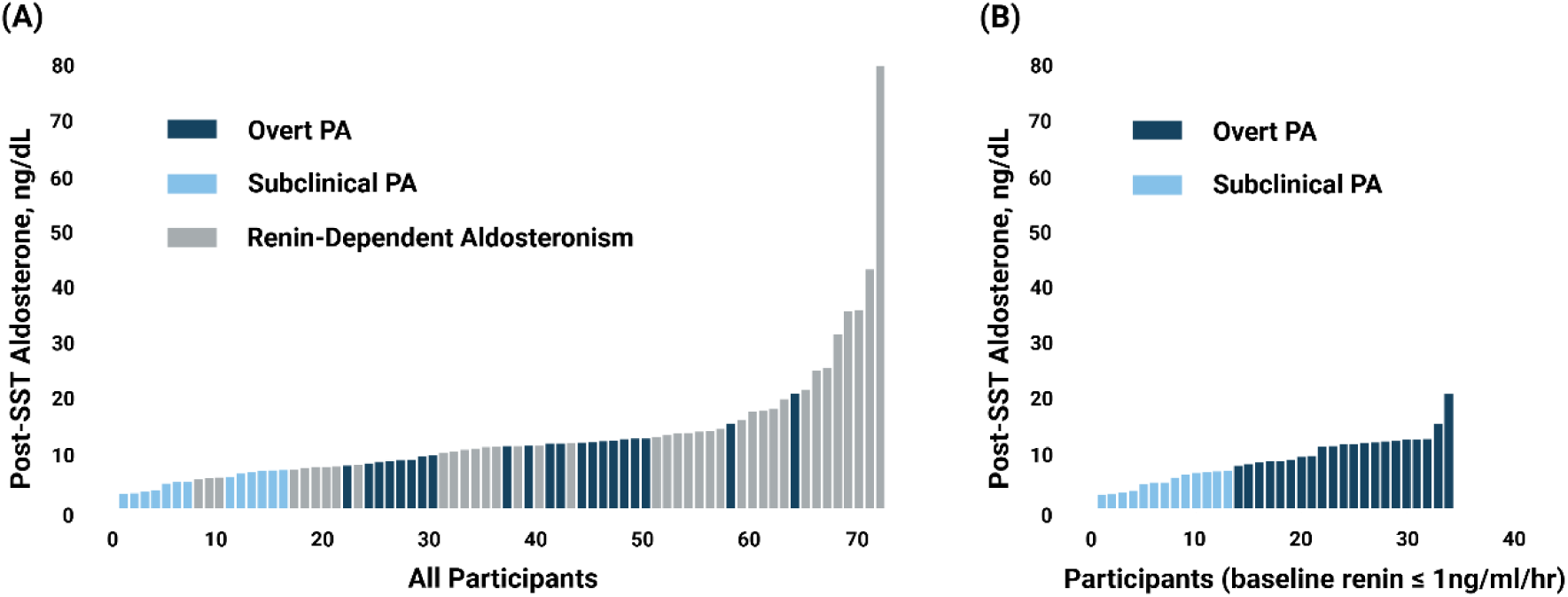
Dysregulated Aldosterone Production in Obese Hypertension. (A) Continuous distribution of post-saline suppression test aldosterone concentrations among all study participants. Overt PA was defined based on the 2025 Endocrine Society diagnostic criteria for primary aldosteronism (baseline PRA ≤ 1 ng/mL/h and post-SST PAC ≥ 7.8 ng/dL by immunoassay). Subclinical PA was defined as baseline PRA ≤ 1 ng/mL/h and post-SST PAC < 7.8 ng/dL, and Renin-Dependent Aldosteronism as baseline PRA > 1ng/mL/h. (B) Distribution of post-saline suppression aldosterone concentrations among participants with baseline plasma renin activity ≤ 1 ng/mL/hr. Individuals meeting the diagnostic criteria for overt primary aldosteronism are marked in dark blue (PRA ≤ 1 ng/mL/h and post-SST PAC ≥ 7.8 ng/dL by immunoassay). The remainder, representing Subclinical PA, are labeled in light blue.

### Cardiac Structural, functional, and cardiometabolic adiposity indicators

The detailed cardiac structural and functional parameters and cardiometabolic adiposity measures are listed in **Table 1**. In this population with zero-to one-drug hypertension and without clinical cardiovascular disease, cardiac structure, function, and myocardial perfusion parameters were largely in the normal range. The mean LV ejection fraction was 59.5 ± 5.6%, the mean LVMI was 49.1 ± 10.6 g/m² and mean LV global longitudinal strain was -18.6 ± 3.4 %. For the cardiometabolic adiposity indicators, the mean total cardiac fat volume was 217.5 ± 75.2 mL, mean visceral-to-subcutaneous fat ratio was 0.9 ± 0.5 and hepatic fat content was 17.2 ± 11.4%.

### Association of post-SST aldosterone excess with cardiac remodeling and adiposity

There was a significant positive association of greater post-SST PAC and greater LVMI, less negative (and thus more abnormal) LV global longitudinal strain, greater cardiac fat volume, and a greater visceral-to-subcutaneous fat ratio. After multivariable adjustment for age, sex, body mass index, 24-hour ambulatory systolic blood pressure, and eGFR, these associations remained statistically significant (**Table 2**). ECV, LV ejection fraction, left atrial parameters, myocardial perfusion reserve, and hepatic fat content were not significantly associated with post-SST PAC. In the sensitivity analysis restricted to participants with complete visceral-to-subcutaneous fat data, the results remained consistent.

**Table 2.** Associations Between Post-SST Aldosterone Levels and Cardiac Structural, Functional, and Adiposity Parameters.

| Log SST Aldosterone | Univariable analysis |  | Multivariable analysis* |  |
| --- | --- | --- | --- | --- |
|  | Beta (95% CI) | *p* value | Beta (95% CI) | *p* value |
| **LV Structure** |  |  |  |  |
| LV mass index, g/m<sup>2</sup> | 0.007 (0.002, 0.013) | 0.014 | 0.016 (0.008, 0.024) | <0.001 |
| LV end-systolic volume index, mL/m<sup>2</sup> | 0.007 (-0.003, 0.016) | 0.176 | 0.005 (-0.006, 0.017) | 0.381 |
| LV end-diastolic volume index, mL/m<sup>2</sup> | 0.004 (-0.001, 0.008) | 0.112 | 0.005 (-0.001, 0.010) | 0.083 |
| LV extracellular volume fraction, % | -1.487 (-4.168, 1.194) | 0.272 | 0.293 (-3.789, 4.375) | 0.886 |
| **LV function** |  |  |  |  |
| LV ejection fraction, % | 0.001 (-0.011, 0.012) | 0.928 | 0.006 (-0.006, 0.018) | 0.341 |
| LV global longitudinal strain, % | 0.019 (0.001, 0.037) | 0.039 | 0.021 (0.001, 0.040) | 0.038 |
| LV global circumferential strain, % | 0.001 (-0.016, 0.018) | 0.919 | 0.002 (-0.018, 0.021) | 0.863 |
| **Left Atrium** |  |  |  |  |
| Left atrial volume index, mL/m<sup>2</sup> | -0.004 (-0.009, 0.001) | 0.136 | -0.004 (-0.009, 0.002) | 0.184 |
| Left atrial ejection fraction, % | -0.001 (-0.007, 0.005) | 0.710 | -0.001 (-0.007, 0.016) | 0.850 |
| Left atrial strain, % | -0.002 (-0.008, 0.004) | 0.506 | -0.001 (-0.008, 0.006) | 0.764 |
| **Cardiac Perfusion** |  |  |  |  |
| Resting Myocardial Blood Flow, mL/min/g | 0.108 (-0.248, 0.464) | 0.547 | 0.280 (-0.143, 0.702) | 0.190 |
| Stress Myocardial Blood Flow, mL/min/g | 0.020 (-0.082, 0.123) | 0.694 | 0.024 (-0.090, 0.139) | 0.676 |
| Myocardial perfusion reserve | 0.001 (-0.096, 0.099) | 0.976 | -0.025 (-0.135, 0.085) | 0.656 |
| **Cardiometabolic adiposity** |  |  |  |  |
| Cardiac fat volume, mL | 0.001 (0.001, 0.002) | 0.022 | 0.001 (0.001, 0.002) | 0.023 |
| Visceral-to-subcutaneous fat ratio | 0.231 (0.110, 0.352) | <0.001 | 0.239 (0.083, 0.396) | 0.003 |
| Hepatic fat, % | 0.149 (-0.455, 0.754) | 0.622 | 0.226 (-0.453, 0.906) | 0.506 |

Visceral-to-subcutaneous fat ratio was measured in 59 of 72 participants, while hepatic fat content was measured in 56 of 72 participants.
\* Models were adjusted for age, sex, body mass index, 24-hour ambulatory systolic blood pressure and estimated glomerular filtration rate. Myocardial perfusion reserve is the unitless ratio of stress divided by rest myocardial blood flow.
**Abbreviations:** LV = left ventricular; SST = saline suppression test.

To investigate whether baseline renin levels influenced the association between post-SST aldosterone production and cardiac structure, function, and cardiometabolic adiposity, we conducted an exploratory analysis incorporating suppressed baseline renin status into the multivariable linear regression model. After adjusting for age, sex, body mass index, 24-hour ambulatory systolic blood pressure, eGFR, and baseline renin status, aldosterone dysregulation remained significantly associated with LV mass index, cardiac fat volume, and visceral-to-subcutaneous fat ratio, but not with LV global longitudinal strain (**Table 3**).

**Table 3.** Exploratory analysis exploring the influence of baseline renin status on the association between dysregulated aldosterone and cardiac structure, function, and cardiometabolic adiposity.

|  | Log SST PAC* |  | Log OSLT PAC* |  | Log DST PAC* |  | Log ACTH stimulation PAC* |  | Global P value* |
| --- | --- | --- | --- | --- | --- | --- | --- | --- | --- |
|  | Beta (95% CI) | p value | Beta (95% CI) | p value | Beta (95% CI) | p value | Beta (95% CI) | p value |  |
| LV mass index | 0.015 (0.007, 0.022) | <0.001 | 0.012 (0.004, 0.020) | 0.003 | 0.015 (0.007, 0.023) | <0.001 | 0.015 (0.008, 0.022) | <0.001 | <0.001 |
| LV global longitudinal strain | 0.015 (-0.004, 0.033) | 0.117 | 0.006 (-0.014, 0.025) | 0.572 | 0.004 (-0.015, 0.024) | 0.008 | 0.005 (-0.013, 0.023) | 0.571 | 0.425 |
| Cardiac fat volume | 0.001 (0.001, 0.002) | 0.041 | 0.001 (0.001, 0.002) | 0.009 | 0.001 (0.001, 0.002) | 0.103 | 0.001 (0.001, 0.002) | 0.093 | 0.014 |
| Visceral-to-subcutaneous fat ratio | 0.218 (0.071, 0.365) | 0.004 | 0.202 (0.050, 0.354) | 0.010 | 0.135 (-0.033, 0.304) | 0.113 | 0.195 (0.042, 0.348) | 0.013 | 0.005 |

Visceral-to-subcutaneous fat ratio was measured in 59 of 72 participants.
\*Individual p values were calculated using multivariable linear regression adjusted for age, sex, body mass index, 24-hour ambulatory systolic blood pressure, estimated glomerular filtration rate, and baseline renin (plasma renin activity $\leq 1$ or $>1$ ). Global p values were derived from multivariable linear mixed-effects models with the same covariate adjustments.
**Abbreviations:** ACTH = adrenocorticotropic hormone; DST = dexamethasone suppression test; LV =left ventricular; OSLT = oral sodium loading test; PAC = plasma aldosterone concentration; SST = saline suppression test.

### Angiotensin II- and ACTH-mediated Aldosterone production is consistently associated with LVMI and cardiometabolic fat

To confirm the consistency of the relationships observed between post-SST aldosterone concentrations and cardiac and adipose parameters, we further examined the associations of these parameters with aldosterone concentrations from three additional physiological phenotyping maneuvers: OSLT, DST, and ACTH stimulation tests, which interrogated angiotensin II-independent, ACTH-independent, and ACTH-dependent aldosteronism, respectively. Individual associations are shown in **Table 4**. After adjusting for age, sex, body mass index, 24-hour ambulatory systolic blood pressure, and eGFR, these repeated aldosterone measurements remained consistently and positively associated with LVMI (global p-value < 0.001), cardiac fat volume (global p-value = 0.009), and the ratio of visceral-to-subcutaneous fat (global p-value = 0.004) (**Figure 4**). A consistent, statistically significant association was not observed between aldosteronism and LV global longitudinal strain across the physiologic phenotyping maneuvers. These results were similar in the exploratory analysis with suppressed baseline renin status included in the multivariable model (**Table 3**).

**Figure 4.**
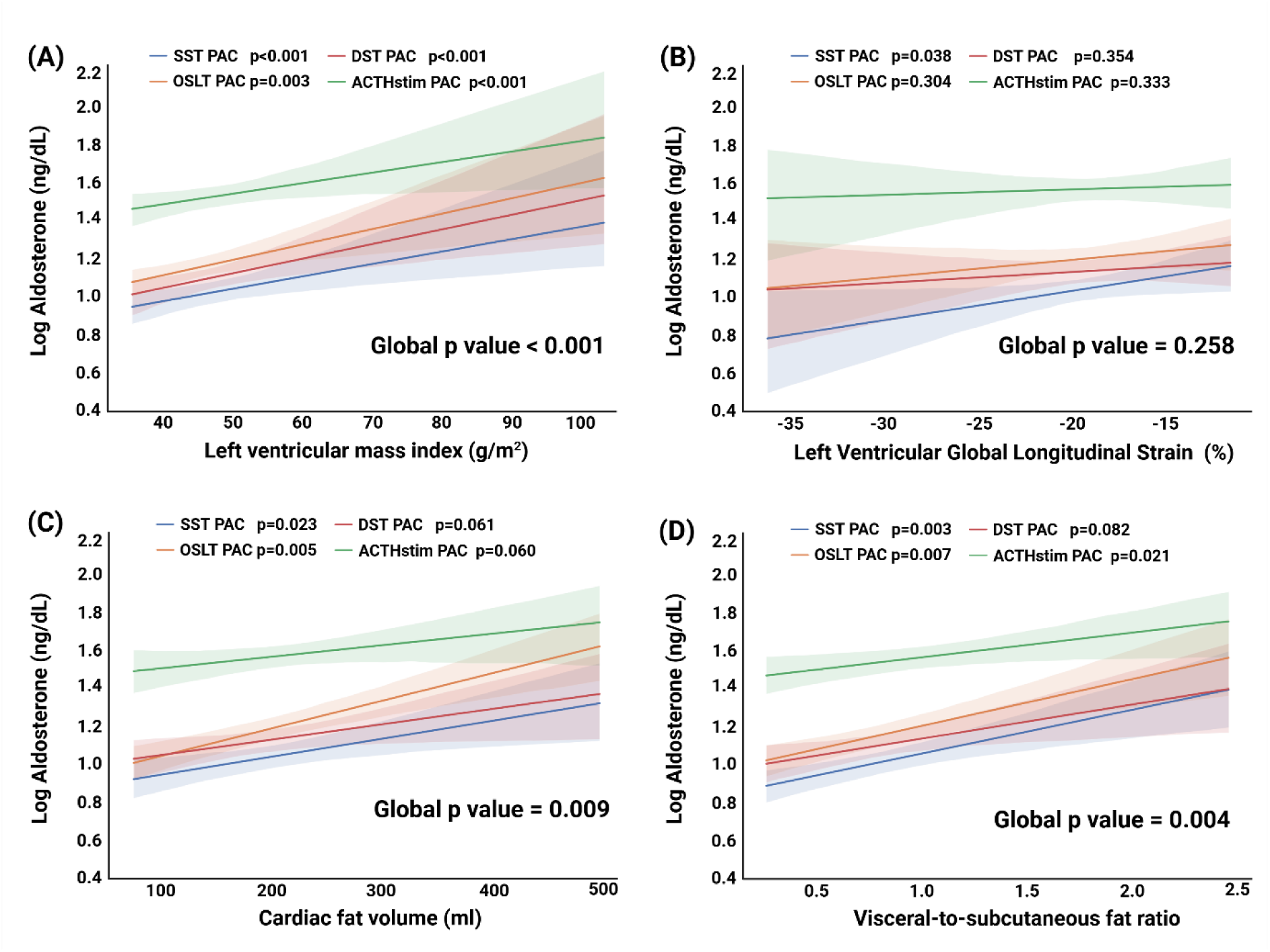
Associations of Aldosterone with Cardiac Structural, Functional, and Adiposity Parameters in Obese Hypertension. Individual p values were calculated using multivariable linear regression adjusted for age, sex, body mass index, 24-hour ambulatory systolic blood pressure, and estimated glomerular filtration rate. Global p values were derived from multivariable linear mixed-effects models with the same covariate adjustments. **Abbreviations:** ACTHstim test = adrenocorticotropic hormone stimulation test; DST = dexamethasone suppression test; PAC = plasma aldosterone concentration; OSLT = oral salt loading test; SST = saline suppression test

**Table 4.** Associations Between Aldosterone and LV mass, LV strain and Adiposity Parameters.

|  | Univariable analysis |  | Multivariable analysis* |  |
| --- | --- | --- | --- | --- |
|  | Beta (95% CI) | *p* value | Beta (95% CI) | *p* value |
| **Log OSLT aldosterone** |  |  |  |  |
| LV mass index, g/m<sup>2</sup> | 0.009 (0.003, 0.014) | 0.003 | 0.013 (0.004, 0.021) | 0.003 |
| LV global longitudinal strain, % | 0.011 (-0.007, 0.029) | 0.229 | 0.010 (-0.010, 0.031) | 0.304 |
| Cardiac fat volume, ml | 0.001 (0.001, 0.002) | <0.001 | 0.001 (0.001, 0.002) | 0.005 |
| Visceral-to-subcutaneous fat ratio | 0.246 (0.123, 0.370) | <0.001 | 0.220 (0.062, 0.378) | 0.007 |
| **Log DST aldosterone** |  |  |  |  |
| LV mass index, g/m<sup>2</sup> | 0.008 (0.002, 0.014) | 0.006 | 0.015 (0.007, 0.023) | <0.001 |
| LV global longitudinal strain, % | 0.007 (-0.011, 0.025) | 0.437 | 0.009 (-0.011, 0.030) | 0.354 |
| Cardiac fat volume, ml | 0.001 (-0.001, 0.002) | 0.051 | 0.001 (-0.001, 0.002) | 0.061 |
| Visceral-to-subcutaneous fat ratio | 0.180 (0.047, 0.313) | 0.009 | 0.153 (-0.020, 0.326) | 0.082 |
| **Log ACTH-stimulated aldosterone** |  |  |  |  |
| LV mass index, g/m<sup>2</sup> | 0.006 (0.001, 0.011) | 0.031 | 0.015 (0.008, 0.022) | <0.001 |
| LV global longitudinal strain, % | 0.005 (-0.012, 0.022) | 0.553 | 0.009 (-0.009, 0.027) | 0.333 |
| Cardiac fat volume, ml | 0.001 (-0.001, 0.001) | 0.110 | 0.001 (-0.001, 0.002) | 0.060 |
| Visceral-to-subcutaneous fat ratio | 0.130 (0.006, 0.254) | 0.040 | 0.207 (0.053, 0.361) | 0.021 |

Visceral-to-subcutaneous fat ratio was measured in 59 of 72 participants.
\* Models were adjusted for age, sex, body mass index, 24-hour ambulatory systolic blood pressure and estimated glomerular filtration rate.
The OSLT was used to interrogate renin- and angiotensin-II independent aldosterone production. The DST and ACTH-stimulation test were used to interrogate ACTH-mediated aldosterone production.
**Abbreviations:** ACTH = adrenocorticotrophic hormone; DST = dexamethasone suppression test; LV = left ventricular; OSLT = oral salt loading test

## Discussion

This study demonstrates that dysregulated aldosterone production and overt primary aldosteronism are prevalent and reproducible phenotypes in a community-dwelling cohort of people with overweight to obesity and mild hypertension. Using modern definitions of primary aldosteronism based on an SST, 29.2% of these participants met criteria for overt primary aldosteronism. Beyond echoing prior studies establishing the high prevalence of primary aldosteronism and dysregulated aldosterone production in hypertension^32–37^, our study delineates the clinical consequences of this aldosterone dysregulation. Using deep phenotyping with controlled physiological testing and high-resolution MRI, we found that dysregulated aldosterone production – including renin-independent, renin-dependent, and ACTH-modifiable aldosteronism – was significantly associated with markers of adverse cardiac remodeling, impaired cardiac function, and increased cardiometabolic fat accumulation, notably within a relatively lower-risk cohort with mild hypertension. The robustness of this association is highlighted by the consistency across four distinct physiological tests examining both renin-angiotensin II and ACTH pathways and by persistence despite multivariable adjustments. Collectively, these findings highlight the role of dysregulated aldosterone production as a distinct and potentially modifiable driver of adverse cardiac remodeling and cardiometabolic fat accumulation in obesity.

Primary aldosteronism exists on a continuum, from normotension to overt disease, rather than as a binary condition^34, 38–41^, and dysregulated aldosterone production across the primary aldosteronism spectrum has been previously linked to cardiovascular remodeling and dysfunction^42–45^. Renin-independent aldosterone production, characteristic of primary aldosteronism, has been associated with left ventricular hypertrophy, metabolic syndrome, impaired global longitudinal strain, and increased cardiometabolic adiposity^44, 46–48^. In this cohort, excess aldosterone production was common, as evidenced by the lack of physiologic suppression following inhibition of the renin-angiotensin system using intravenous saline infusion. Further, we here show that similar pathologic associations with cardiac remodeling and adiposity are present in individuals with obesity and mild hypertension, even in the absence of clinically diagnosed primary aldosteronism. This underscores the need to recognize dysregulated aldosterone production as a prevalent and potentially modifiable contributor to cardiometabolic risk in obese individuals, even at early stages of hypertension.

While obesity is increasingly recognized as a contributor to aldosteronism, its underlying mechanisms are complex^6, 7^ and likely involve both renin-independent and renin-dependent pathways^6, 7, 49, 50^, with both phenotypes prevalent in our study. Adipose tissue, particularly cardiometabolic fat, acts as an active endocrine organ that stimulates aldosterone production. Elevated leptin and very low-density lipids levels have been shown to upregulate CYP11B2 expression in adrenal cells, enhancing aldosterone synthesis^15^. Furthermore, studies demonstrated that complement-C1q TNF-related protein 1 (CTRP1), expressed at high levels in adipose tissue and the adrenal zona glomerulosa, is a potent aldosterone-stimulating factor^51^. Human adipocytes also secrete mineralocorticoid-releasing factors that further potentiate adrenal aldosterone production^13^. Altered ACTH dynamics in obesity may also contribute to renin-independent aldosterone production^52^.

Obesity may also promote aldosterone production through renin-dependent pathways mediated by heightened sympathetic nervous system activity and impaired responses to expanded intravascular volume^49, 50^. Consistent with this, our exploratory analysis demonstrated that aldosterone dysregulation was associated with adverse cardiac structure, function, and adiposity, regardless of baseline renin status. This supports the concept that multiple, overlapping mechanisms of aldosterone excess and mineralocorticoid receptor activation can coexist in obesity. In addition, the complex hormonal and hemodynamic changes associated with obesity can obscure the diagnosis of primary aldosteronism, leading to its substantial under-recognition in this population^53^.

Mechanistic studies have implicated aldosterone-mediated mineralocorticoid receptor activation as a pathologic driver in both adipose and cardiac tissues, and dysregulated aldosterone production has been implicated in adverse cardiovascular outcomes^8, 54, 55^, including in mediating cardiovascular risk in obesity^6, 56, 57^. In adipose tissue, mineralocorticoid receptor signaling promotes adipocyte hypertrophy, inflammation, and insulin resistance^58–60^. In the myocardium, aldosterone contributes to cardiac hypertrophy, macrophage infiltration, interstitial fibrosis, and cardiac dysfunction^55, 61–63^. Aligned with our findings, Shah et al. demonstrated that pericardial fat, but not hepatic fat, is associated with cardiovascular outcomes and cardiac remodeling^27^. Our findings further strengthen these observations, suggesting that dysregulated aldosterone-MR signaling may represent a mechanistic link between adiposity and subclinical cardiac remodeling, while acknowledging the potential for bidirectional interactions between adipose and cardiovascular tissues. More significantly, we extend observations from large imaging cohorts by implicating excess, non-suppressible aldosterone production as a potential upstream driver of cardiovascular and metabolic risks associated with visceral and cardiac fat in obese hypertensive individuals^27, 28, 64, 65^. This supports the concept that aldosterone excess in obesity activates a unique pathophysiological pathway, the adipose-aldosterone interaction, which may lead to detrimental cardiometabolic outcomes through mineralocorticoid receptor activation. Consequently, these data provide rationale for expanding the consideration of aldosterone-targeted interventions in obesity. Mineralocorticoid receptor antagonists and aldosterone synthase inhibitors may hold potential to reverse early remodeling associated with dysregulated aldosteronism, independent of weight or blood pressure reduction.

The interpretation of our findings should consider several limitations. First, our study population consisted exclusively of overweight and obese individuals with stage 1–2 hypertension, which may limit the generalizability of our findings. However, by enrolling participants with mild, minimally treated hypertension, our study provides unique insight into obesity related aldosterone dysregulation. Second, we did not directly assess tissue-level mineralocorticoid receptor activation or fibrotic or inflammatory biomarkers, which could have provided additional mechanistic insights into the pathophysiology of dysfunctional adiposity. Third, while comprehensive cardiac and abdominal MRI was performed, not all participants had complete measurements for hepatic fat content or visceral-to-subcutaneous fat ratio due to inadequate imaging quality in some cases. This might introduce a minor selection bias and potentially impact the generalizability of these specific adiposity findings, though image availability was unrelated to aldosterone physiology. However, the associations with aldosterone remained consistent in a sensitivity analysis in those without missing data. Finally, the cross-sectional design limits causal inference. Although MRI allowed for precise quantification of fat distribution and cardiac remodeling, longitudinal imaging studies are needed to determine the temporal effects of aldosterone dysregulation and the impact of aldosterone-targeted therapies including mineralocorticoid receptor antagonists and the emerging aldosterone synthase inhibitors^66^.

## Conclusion

In this study of obese adults with mild hypertension, we identified a prevalent and reproducible phenotype of dysregulated aldosterone production, and frequent overt primary aldosteronism, that is independently associated with adverse cardiac remodeling and increased cardiometabolic adiposity. These findings implicate a pathophysiologic adipose–aldosterone interaction and identify dysregulated aldosterone production as an under-recognized but potentially modifiable hormonal driver of obesity-related cardiovascular risk. Future studies are warranted to determine whether targeted suppression of aldosterone and mineralocorticoid receptor activation can mitigate early cardiometabolic injury in this growing population.

